# The cross-site reproducibility of MRI morphometric phenotypes in psychiatric disorders

**DOI:** 10.1101/2025.07.09.25331220

**Authors:** Trang Cao, James C. Pang, Mehul Gajwani, Ashlea Segal, Alexander Holmes, Joshua F Wiley, Sidhant Chopra, Juan Helen Zhou, Christopher LH Chen, Fang Ji, Ben J Harrison, Christopher G Davey, Toby Constable, Jeggan Tiego, Bree Hartshorn, Jessica Kwee, Mark A. Bellgrove, Alex Fornito

## Abstract

Decades of structural magnetic resonance imaging (MRI) studies have documented alterations of grey matter morphometry in psychiatric disorders, but the field has failed to identify any consensus disease phenotypes. Here, we examine whether current approaches will ever converge on such phenotypes by evaluating the consistency of brain-wide maps of grey matter volume and cortical thickness differences obtained for each of 59 study sites of five psychiatric disorders (schizophrenia, schizoaffective disorder, autism spectrum disorder, major depressive disorder, and bipolar disorder), totaling 2437 patients and 2065 controls. We find that cross-site consistency is low (median r ≤ 0.16); markedly reduced compared to Alzheimer’s Disease (r = 0.54); unexplained by demographic, clinical, or scanner differences; and robust to analytic choices. Using bootstrapping, we observe that consistency may improve for sample sizes ≥200 per group for schizophrenia, but that other disorders may require much larger samples. Our findings indicate that current widespread practices in structural MRI are unlikely to identify robust morphometric phenotypes for psychiatric disorders.

## Introduction

Over a century ago, Kraepelin established the modern descriptive approach to psychiatric diagnosis by using symptomatologic and prognostic features to discriminate between dementia praecox and manic-depressive illness^1^. He also proposed that distinct psychiatric disorders should be associated with clear and diagnostically specific pathophysiological changes in the brain, an idea that has underpinned the majority of subsequent biological psychiatric research. The advent of magnetic resonance imaging (MRI) in the 1980s afforded an unprecedented capacity to map brain structure and function in living patients, raising hopes that such core neurobiological phenotypes of disease would be identified. T1-weighted structural MRI has arguably been the most widely used tool to interrogate grey matter correlates of mental illness, with decades of work and thousands of studies reporting differences in grey matter morphometric measures, such as volume and cortical thickness, between patients and controls. However, despite this voluminous literature, we still lack widely-accepted, clinically actionable morphometric (e.g., cortical thickness and/or grey matter volume) phenotypes that can inform diagnosis, treatment response, or monitoring of risk and progression for psychiatric disorders^2^. Instead, the literature is littered with findings that are inconsistent with respect to the specific brain regions implicated in a given disorder and the polarity of case-control differences, even within the same region. For example, both increased and decreased grey matter volume and/or cortical thickness have been reported in the striatum of people with schizophrenia^3^; in the superior temporal sulcus and the frontal lobe in association with autism^4–7^; in the orbitofrontal cortex, parietal cortex, temporal cortex, occipital cortex, insula^8^, rostral anterior cingulate^8–10^, pericalcarine^9,11^, and thalamus^9,12^ in major depression; and in the anterior cingulate^13–16^ and ventral prefrontal cortex ^13,17^ in bipolar disorder.

There are multiple potential reasons for such inconsistencies, including variations between studies with respect to the clinical and demographic characteristics of the participants (e.g., illness severity/duration, age of onset, medication exposure, and substance use); variations in the image processing and analysis techniques used^18–20^; a traditional reliance on small samples, which can yield effects with wide confidence intervals that are highly prone to sampling variability^21,22^; and the use of classical case-control designs relying on group-mean comparisons, which entail multiple assumptions that may be insufficient to parse the inherent heterogeneity of psychiatric disorders (for an overview, see ^2^). Additional, more fundamental challenges are posed by the limited resolution and mechanistic specificity of MRI, as well as current, symptom-based nosological systems, such as the Diagnostic and Statistical Manual of Mental Disorders (DSM)^23^, which generally show limited biological validity ^24,25^, within-construct homogeneity^26,27^ (i.e., diagnostic criteria can often be met through myriad symptom combinations^28–30^), and diagnostic specificity^31,32^. Averaging across individuals defined using such constructs may yield group means that do not adequately represent individual cases^33–36^, driving inconsistencies across studies.

These considerations raise the fundamental question of whether current practices in structural MRI studies can ever converge on a robust morphometric phenotype for psychiatric disorders, as defined using current diagnostic systems. To address this question, we collated participant-level data on 2437 people diagnosed with one of five psychiatric disorders––schizophrenia (SCZ), schizoaffective disorder (SCA), autism spectrum disorder (ASD), major depressive disorder (MDD), or bipolar disorder (BD)––and 2065 controls acquired across 59 distinct study sites to assess the cross-site consistency of grey matter morphometry differences in each condition and to identify potential factors that may influence such consistency. We benchmarked our consistency estimates against an additional seven sites comprising scans in 654 people with Alzheimer’s disease (AD) and 937 controls, given that AD is associated with a well-described neurodegenerative phenotype^37^. Our analysis shows that grey matter differences mapped for each psychiatric disorder, estimated either with respect to cortical thickness or regional grey matter volume, generally show low consistency across different studies. We identify the specific factors that may influence the variations in cross-site consistency and provides estimates of the sample sizes required to improve the robustness of findings in the literature.

## Results

### Analysis workflow for evaluating cross-site consistency of grey matter differences

We collated individual-level data from 25 datasets acquired across 59 sites with T1-weighted MRI in people with psychiatric disorders and healthy controls (see Supplementary Table 1 for a summary and 1 in Online Methods). We also aggregated data on AD from an additional 5 datasets collected across 7 sites. We only included a site if it contained data for at least 20 participants per group––i.e., controls and cases––from the same scanner after quality control (see 2 in Online Methods). All participants in the psychiatric studies were aged between 18 and 60 years. For the AD studies, participants were aged between 42 and 95 years. Summaries of the data available in each disorder are presented in Table 1 and Supplementary Table 2, with site-specific demographics provided in Supplementary Tables 3 and 4. MRI detailed are in Supplementary Table 5.

**Table 1.**
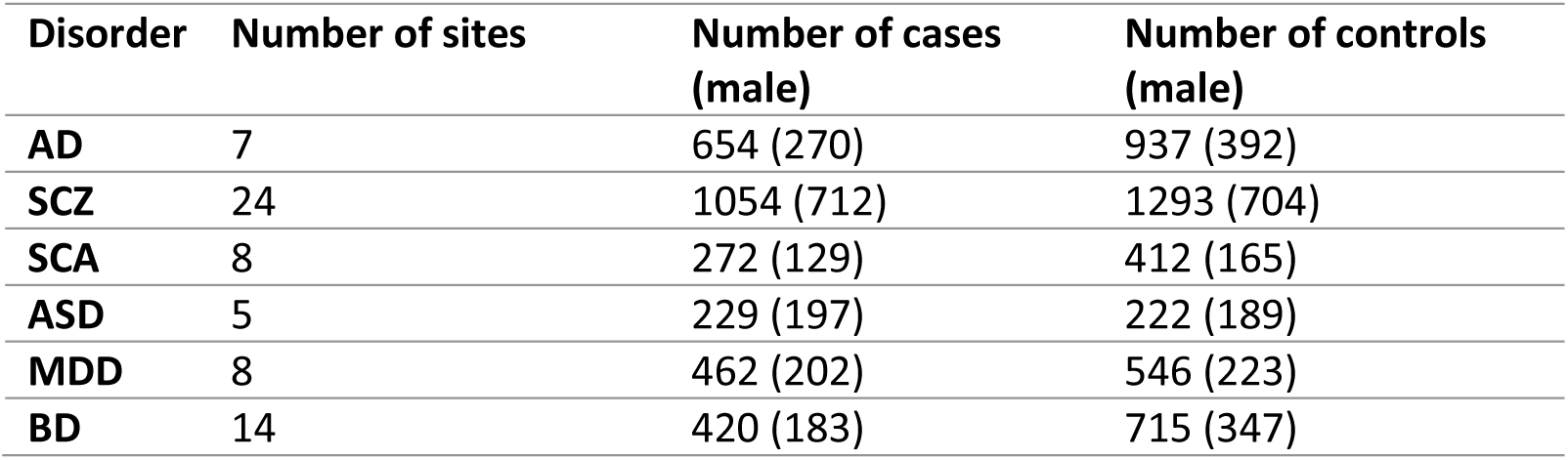
Summary of data for each disorder in the SBM analysis. Note that some sites have multiple disorders and the controls are the same for different disorders in these sites.

We applied uniform image processing protocols on data from all sites for surface-based morphometry (SBM), which was used to estimate cortical thickness (using FreeSurfer^38,39^), and voxel-based morphometry (VBM), which was used to estimate grey matter volume (using SPM^40^) (see 3 in Online Methods). Statistical comparisons between cases and controls were performed using general linear models with covariates (age and sex for SBM and age, sex, and total intracranial volume for VBM, as commonly considered^41^) independently at each site, resulting in a site-specific *z*-statistic map (for SBM) or *t*-statistic map (for VBM) that quantifies group mean differences in cortical thickness or grey matter volume in each cortical surface vertex or brain voxel, respectively (Fig. 1). For each disorder, cross-site consistency (CSC) was quantified using product-moment spatial correlations between each pair of site-specific maps (Fig. 1; see 4 in Online Methods). To verify the statistical significance of the consistency estimates, we compared the observed values to null models obtained by correlating randomized maps with similar autocorrelation to the empirical data as generated using eigenstrapping^42^ and BrainSMASH^43^. We also considered a permutation-based null model that involved shuffling patient and control labels before re-estimating group difference maps^44,45^ (see 5 in Online Methods). Note that our use of a unified image processing and analysis pipeline approximates an upper bound on the consistency that can be achieved in the literature, given that, in reality, different investigators make different processing and analysis choices, which likely compounds the degree of inconsistency between studies.

**Fig. 1.**
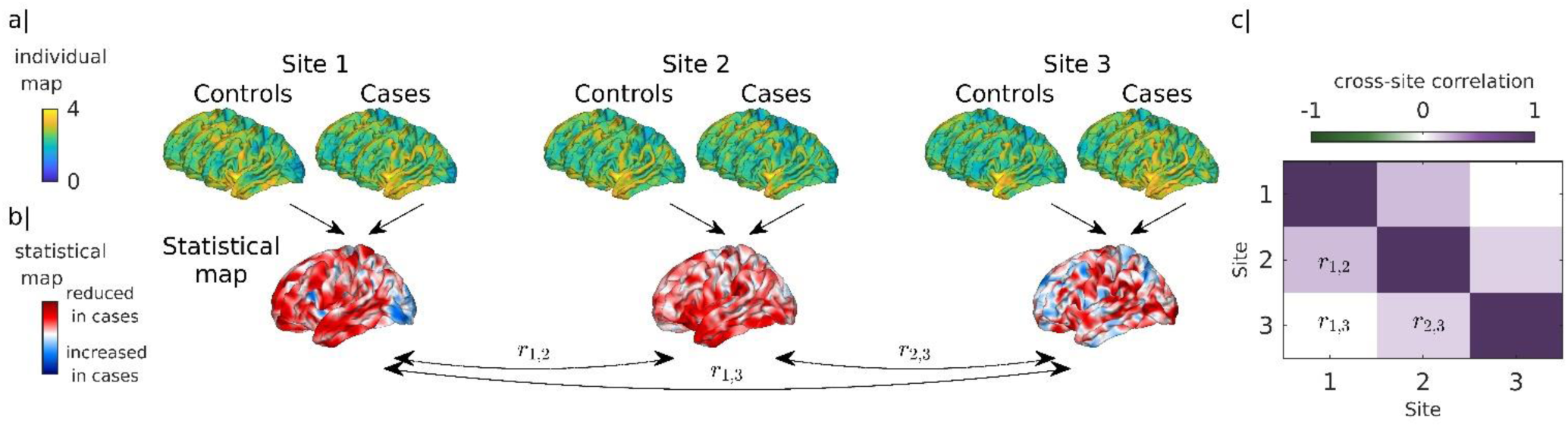
Analysis pipeline. (a) Each site comprises a group of controls and cases. We obtained maps of cortical thickness (shown here) or grey matter volume (not shown) for each individual in each group at each site. (b) For each site, we used a general linear model to map group differences at each of thousands of brain locations, resulting in a map of *z*-statistics (for cortical thickness) or *t*-statistics (for volume) quantifying the magnitude of these differences. These maps were then correlated to obtain an estimate of cross-site consistency (CSC) between each pair of sites (e.g., *r*_1,2_ represents the spatial correlation, or CSC, between difference maps for sites 1 and 2). (c) The CSC estimates for every pair of sites can be represented as a symmetric site-by-site CSC matrix for each disorder.

### Cross-site consistency of cortical thickness differences

We first benchmarked our expectations for the CSC of grey matter differences with respect to AD, which has a well-described pathophysiological basis and neurodegenerative phenotype^37^. Fig. 2a shows that, across seven different AD sites, the CSCs for cortical thickness differences in the left hemisphere range between 0.43 ≤ *r* ≤ 0.71 with a median of 0.54 (similar results were obtained for the right hemisphere; see Supplementary Fig. 1).

**Fig. 2.**
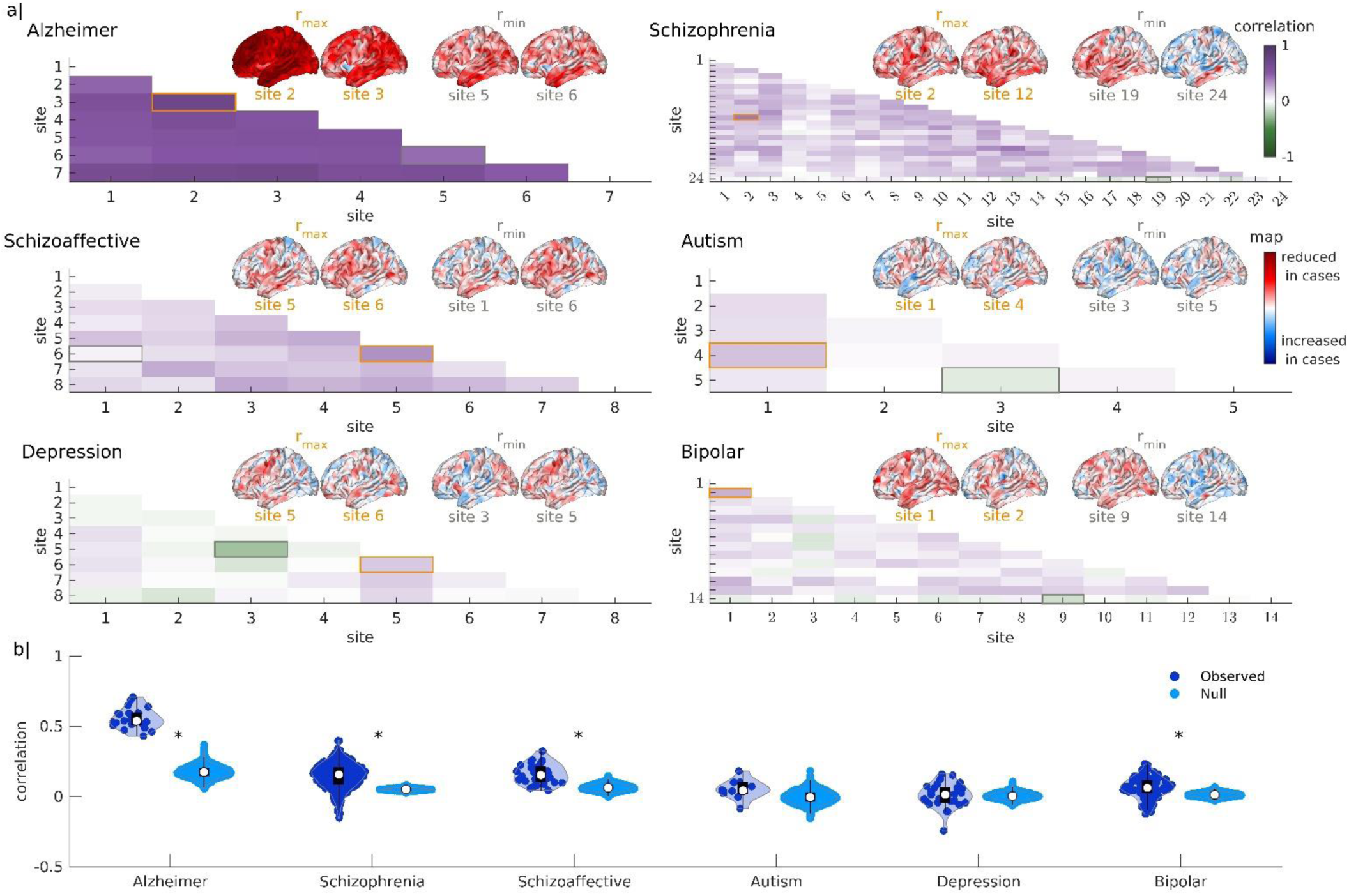
Cross-site consistency (CSC) of site-specific left hemisphere *z*-maps quantified with SBM. (a) Pair-wise CSC estimates for site-specific *z*-maps of the left hemisphere for each disorder. Pairs of maps with the highest CSC (orange box) and the lowest correlation (grey box) are shown for each disorder. The site names corresponding to the site indices are listed in Supplementary Table 3. (b) Distributions of pair-wise CSCs between observed site-specific *z*-maps, along with the corresponding distributions of the median CSCs obtained using a spatially autocorrelated null model^42^. White circles represent the median, black boxes show the interquartile range (IQR), and whiskers extend to the last non-outlier data points within ±1.5×IQR. Light blue outlines the shape of the estimated CSC distribution and dark blue dots represent observed CSC values. Asterisks mark the disorders where the observed median CSC is significantly different from the null distribution (non-parametric two-sided statistical test adjusted for multiple comparisons, *n* = 1000 null, *p* = 0, 0, 0, 0.003 < 0.05).

Repeating the analysis for the five psychiatric disorders, all CSC estimates were lower than those observed in AD (Fig. 2a). SCZ and SCA showed the highest CSC, with estimates ranging between -0.15 ≤ *r* ≤ 0.40 (median = 0.16) and 0.04 ≤ *r* ≤ 0.32 (median = 0.15), respectively. Generally, lower consistency was observed for ASD (-0.09 ≤ r ≤ 0.18) (median = 0.04), MDD (-0.24 ≤ *r* ≤ 0.16) (median = 0.01), and BD (-0.13 ≤ *r* ≤ 0.23) (median = 0.06). Fig. 2b shows that the median CSC was significantly greater than the null expectations (*p* < 0.05) for all disorders except for ASD and MDD. Similar results were obtained for both left and right hemispheres (Extended Data Fig. 1), when using VBM instead of SBM (Extended Data Fig. 2), and when using different nulls (Supplementary Fig. 1 and 2). Together, these analyses provide limited support for grey matter thickness or volume differences, as commonly investigated in structural MRI studies, as representing robust morphometric phenotypes in psychiatric disorders, standing in stark contrast to the findings for AD. Our findings further suggest that the levels of consistency observed for ASD and MDD are consistent with random expectations.

### Effects of data processing and analysis pipelines on cross-site consistency

We initially quantified CSC estimates between pairs of unthresholded difference maps, the analysis of which is proposed to yield more transparent, reproducible results^46^. Most published studies generally use cluster-level statistics to threshold these maps and reveal areas where the group differences exceed some criterion for statistical significance^47,48^. To assess whether this common practice of thresholding influences CSC estimates, we thresholded each contrast map (controls > cases or controls < cases) obtained for each site using *F*-tests in SBM or *t*-tests in VBM using two thresholds: (1) *p* ≤ 0.05, uncorrected; and (2) after cluster-based correction for multiple comparisons, ensuring that any supra-threshold clusters were significant at a familywise error-corrected level of *p* ≤ 0.05 (see 3 in Online Methods). The thresholded maps were then binarized and CSC estimates were quantified using binary correlations^49,50^. We additionally evaluated the replication rate, which quantifies the repetition of significant results (i.e., voxels, vertices, or regions) of one map in other maps, as per prior work^21,51^(see 4 in Online Methods). The impact of thresholding on CSC estimates for SBM and VBM is shown in Extended Data Fig. 3 and Supplementary Fig. 3, respectively. This analysis revealed that CSCs between thresholded maps were even lower than those obtained with unthresholded maps, with most falling within null expectations, especially following multiple comparison correction. These findings indicate that thresholding does not facilitate the identification of robust grey matter thickness or volume differences measured by structural MRI in psychiatric disorders.

Spatial smoothing is another common procedure applied in SBM and VBM to boost the signal-to-noise ratio and render the data more amenable to certain parametric statistical tests. Our initial analyses employed minimal smoothing (e.g., the results in Fig. 2 and Extended Data Fig. 2 were generated with a kernel of 10 and 6 mm full-width at half-maximum, FWHM, respectively) but many studies use much higher levels of smoothing. We therefore re-estimated each site-specific unthresholded and thresholded difference map after smoothing each person’s thickness or grey matter volume maps with smoothing kernels of 15 mm and 20 mm FWHM for SBM and kernels of 8 mm and 12 mm FWHM for VBM. Results obtained with different smoothing kernels for SBM and VBM are presented in Extended Data Fig. 4 and Supplementary Fig. 4. Increased smoothing marginally improved the median CSC for unthresholded maps. For example, the median correlation in schizophrenia increased from 0.16 with 10 mm smoothing to 0.22 with 20 mm smoothing in SBM, and from 0.18 with 6 mm smoothing to 0.20 with 12 mm smoothing in VBM. However, the variance of CSCs also increased with more smoothing (Extended Data Fig. 4 and Supplementary Fig. 4), suggesting that smoothing shifts the distribution of correlations upwards but also accentuates between-site variability. Smoothing did not significantly improve the consistency of thresholded maps (Supplementary Figs. 5 and 6).

Some studies perform analyses at the level of parcellated regions rather than individual voxels or vertices. We therefore parcellated each participant’s cortical thickness map and grey matter volume map using the Desikan-Killiany (DK) (34 regions per hemisphere)^52^ and Schaefer (50, 250, and 500 regions per hemisphere) atlases^53^, which are among the most widely used in the field. For each atlas, we ran separate general linear models (GLMs) in each of the regions. We then estimated the correlations between unthresholded maps, as well as binary correlations and replication rates for thresholded maps. The use of parcellated maps increased CSC estimates slightly, but by no more than 10%, on average. Higher CSC estimates were observed for lower-resolution parcellations (Extended Data Fig. 5 and Supplementary Fig. 7) but, much like the effect of spatial smoothing, the variance of CSC estimates also increased (Supplementary Figs. 5 and 6). We also compared the results when including mean CT as a covariate in SBM analyses and observed a slight decrease in the CSCs (Supplementary Fig. 10).

Prior to each of the above analyses, we adjusted each participant’s cortical thickness and grey matter volume maps for site-specific effects using the commonly used data harmonization method, ComBat^54^. This step may lead to optimistic CSC estimates given that different studies in the field generally do not apply this step with respect to each other. We therefore repeated the analysis without applying ComBat. Repeating the analysis without prior application of ComBat^54^ did not have a noticeable impact on CSC estimates (Extended Data Fig. 6 and Supplementary Fig. 8), indicating that our use of this procedure did not inflate cross-site correlations.

Collectively, these findings indicate that major data processing and analysis choices, such as thresholding strategy, smoothing kernel size, parcellation, and data harmonization, exert only a minor influence on the cross-site consistency of grey matter thickness or volume differences measured by structural MRI.

### Effects of clinical, demographic, and MRI features on cross-site consistency

To evaluate how sample and MRI data features affected estimates of cross-site consistency, we quantified the level of similarity between each pair of sites across 19 different variables related to demographics, clinical information, and image quality and acquisition (see 6 in Online Methods; for a full list of features, see Table 2). This procedure resulted in a site × site matrix for each feature that quantifies the degree of similarity between each pair of sites. We call this matrix the cross-site feature similarity matrix. For each disorder and each characteristic, we extracted the non-redundant elements (i.e., the off-diagonal upper triangle) of the feature similarity matrix and correlated them with the corresponding elements of the CSC matrix (e.g., the matrices depicted in Fig. 2a), allowing us to examine whether CSC estimates track cross-site similarities in different demographic, clinical or scanner features. We used Mantel tests^55^ to determine the statistical significance of these correlations.

**Table 2.** List of sample and data characteristics.

| Sample and data characteristics of each site | Short name for figures |
| --- | --- |
| mean of participant ages | mean age |
| variance of participant ages | var age |
| number of male participants | male |
| number of female participants | female |
| male-to-female ratio of participants | sex ratio |
| number of cases | cases |
| number of controls | controls |
| total number of participants | subjects |
| case-to-control ratio | case control ratio |
| medication exposure | treatment |
| mean Euler number<sup>38</sup> of cortical surface reconstructions | mean EN |
| variance of surface Euler numbers | var EN |
| mean of age of illness onset | mean age onset |
| variance in the age of illness onset | var age onset |
| mean of illness duration | mean illness duration |
| variance of illness duration | var illness duration |
| MRI scanner manufacturer | scanner brand |
| MRI scanner model | scanner model |
| MRI voxel size | voxel size |

Fig. 3 shows the correlations between the independent elements of the SBM CSC matrix and matrix of cross-site feature similarities for each disorder and each feature (see Table 2 for a full list of features). These correlations spanned the range −1 < *r* ≤ 0.6 across disorders and features, with a median of 0. Of the 19 associations tested for each disorder, 2 were statistically significant after Bonferroni correction for multiple comparison. These significant correlations indicated that higher cross-site consistency was associated with lower cross-site similarity in the number of female participants and the sample sex-ratio in autism spectrum disorder. However, only three sites had female participants, meaning that this result should be treated with caution until replicated in larger, more diverse samples. No significant associations were observed between VBM CSC estimates and any of the 19 clinical, demographic, or MRI features across any disorder (Extended Data Fig. 7).

**Fig. 3.**
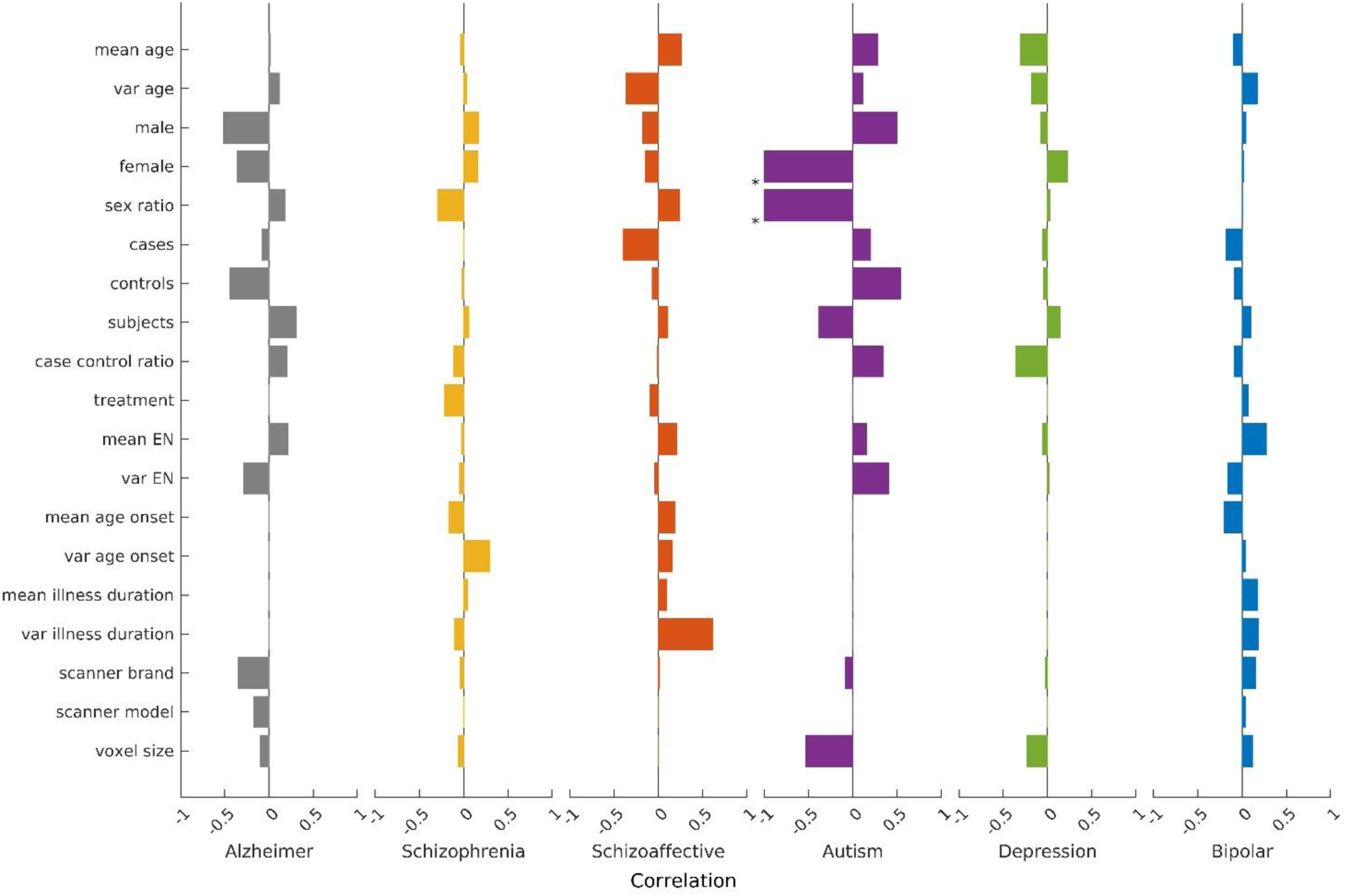
Associations between cross-site consistency estimates and sample/MRI data characteristics. Bars represent correlations between each pair of SBM CSC and cross-site feature similarity estimates for 19 features in each disorder. The asterisks indicate significant associations under the Mantel test^55^ (*n* = 5000 iterations, *p* = 10^−13^, 10^−13^ < 0.05) with Bonferroni correction. See Table 2 for more information on each of the sample and MRI data variables.

### Effects of sample size on cross-site consistency

Single-site psychiatric neuroimaging studies typically examine samples with ∼40 participants per group^56^. Supplementary Table 3 indicates that the smallest sample size observed across the 59 sites considered here is 21 cases and 21 controls, whereas the largest was 115 cases and 145 controls. The median number of participants across all 59 sites was 38 for patients and 49 for controls. Small samples generally yield unreliable effect size estimates^21,57^, with some analyses indicating that 550 samples may be required to detect reliable differences between people with schizophrenia and controls when considering 500 brain regions^51^.

To investigate the influence of sample size on CSC, we used a bootstrapping approach (see 7 in Online Methods). For each disorder, we combined all data from cases and controls to form two populations. From the case population and control population, we randomly sampled two pseudo-sites, each of which comprises a case group and a control group with a predetermined sample size. We chose sample sizes that are approximately equally spaced on a logarithmic scale; i.e., *N* = {10, 16, 25, 40, 63, 100, 158, 251, 398}. According to our data pool, the maximum possible sample sizes (*N*_*max*_) are: 327 for AD, 527 for SCZ, 136 for SCA, 111 for ASD, 231 for MDD, and 210 for BD.

We then computed two *z*-maps of cortical thickness differences––one for each pseudo-site––and correlated these two maps. We repeated the sampling procedure 100 times with replacement to obtain a distribution of such correlations at each sample size (Fig. 4a). We treated the mean bootstrapped-correlations with the maximum possible sample size, denoted 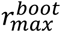, as the most robust correlation estimate that we can obtain for a disorder given our data, as this value corresponds to the CSC obtained with the largest possible data split. Values of 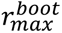 near zero suggest that there is no consistent grey matter thickness or volume differences measured by structural MRI in the disorder that can be identified using the maximum sample sizes considered here, whereas values of 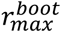 close to one indicate that a reliable phenotype can be identified with a sufficiently large sample size. Repeating the bootstrapping procedure across different sample sizes allowed us to evaluate how quickly the cross-site correlations converge on 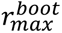 for each disorder. Disorders with a faster convergence rate are associated with a more robust morphometric phenotype. We ran the same bootstrapping procedure with binary correlations and replication rates estimated for thresholded maps. We also ran the procedure on cortical thickness maps parcellated with the DK, Schaefer-100, Schaefer-500, and Schaefer-1000 atlases.

**Fig. 4.**
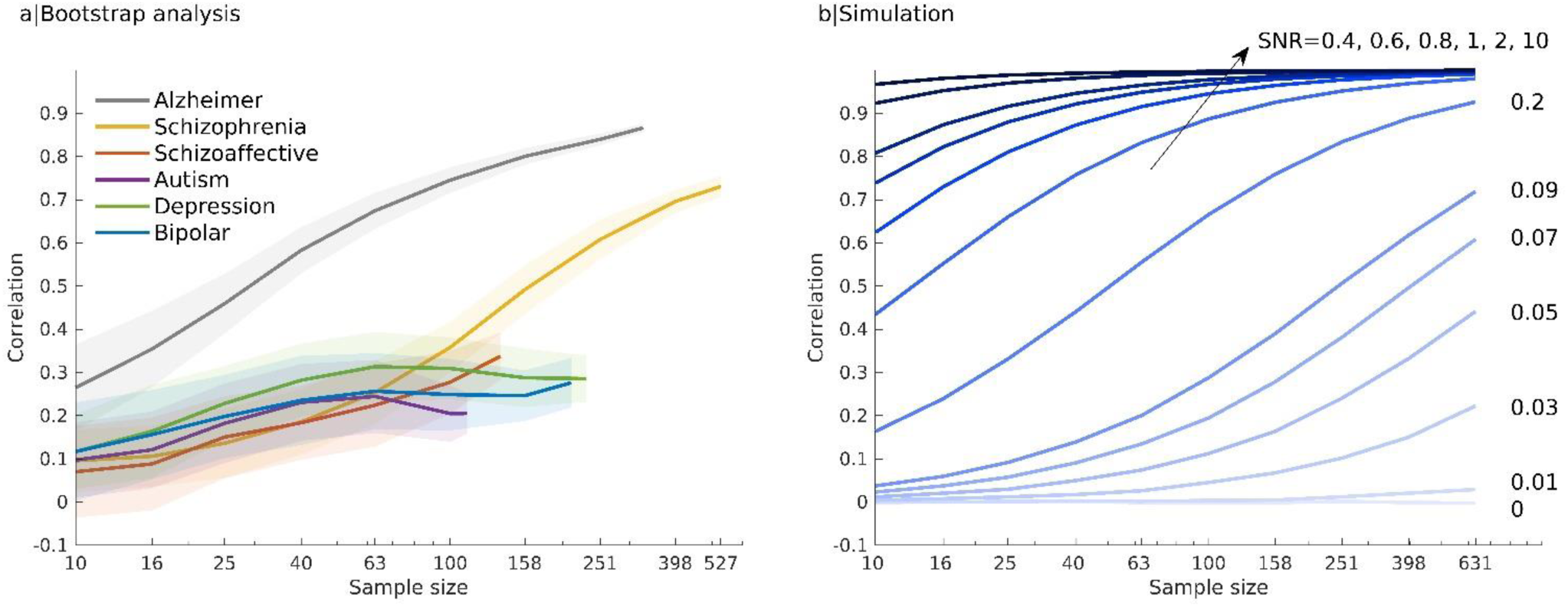
Effects of sample size on cross-site consistency. Mean CSC estimates for vertex-level *z*-maps of pseudo-sites of varying sample size (*x*-axis). (a) Consistency measures from bootstrapped empirical data in each disorder. (b) CSC estimates from a simulation with 100000 vertices for different SNR. Shaded areas represent one standard deviation from the mean.

The results of our bootstrapping analysis are shown in Fig. 4a. The mean bootstrapped-correlations obtained with the maximum possible sample size, 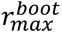, varied considerably across psychiatric disorders, with the highest observed for SCZ (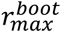 = 0.73) and lowest for ASD (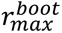 = 0.2). For comparison, 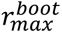 = 0.87 for AD. We additionally found that vertex-level CSC, regardless of whether it is measured by correlation, binary correlation, or replication rate, increases with sample size for AD, SCZ, and SCA (Fig. 4a and Extended Data Fig. 8). In contrast, sample size increases produced only marginal gains for ASD, MDD, and BD.

The analysis of 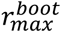 suggests that grey matter thickness or volume differences measured by structural MRI can be identified as robust phenotypes only for AD and schizophrenia-spectrum disorders. However, the maximum sample size available for these disorders, and thus for the estimation of 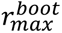, was larger than for the others. Fig. 4a shows that the median bootstrapped consistency at the highest sample size common to all groups (i.e., *N* = 100) is still very high for AD (*r* = 0.75) and higher for schizophrenia (*r* = 0.36) than the other conditions (all *r* < 0.31). SCZ is associated with a dramatic increase in consistency beyond a sample size of 100. SCA showed an increasing trend that tracked the schizophrenia curve, whereas the curves for depression and bipolar disorder appeared flat; for instance, moving from a sample of *N* = 10 to *N* = 158 yields a 10-fold increase in average CSC for schizophrenia compared to a 2-fold gain in MDD. Similar results were obtained for thresholded maps (Extended Data Fig. 8) and when using atlas-parcellated measures (Extended Data Fig. 9). These findings indicate that CSC starts to increase rapidly for sample sizes of *N* > 150 in SCZ, reaching a median of *r* = 0.5 at *N*∼200. SCA may follow a similar trend. Comparable increases in other disorders were not observed, but we had limited data for sample sizes exceeding *N* = 150 in ASD, MDD, and BD.

Given the limitations on the maximum sample sizes available in our data, we complemented these bootstrapping analyses with a simple simulation to evaluate, in a more general sense, how CSCs vary as a function of sample size given assumptions about the degree to which different individuals within the patient group express a common disease phenotype relative to intrinsic inter-individual variability and/or measurement noise (see 8 in Online Methods). The model thus allows us to tune the signal-to-noise ratio (SNR) of the disease phenotype. Fig. 4b shows that, under this model, sample size generally has little impact on CSC estimates when disease phenotype SNR is either very low (e.g., SNR=0.01) or very high (SNR=1). In the former case, CSCs remain low even at large sample size; in the latter, CSCs hit ceiling at small sample sizes (e.g., *N* = 10). For moderate SNR∼0.2, average CSCs can be improved from 0.2 to 0.8 by increasing the sample size from 10 to 250. Comparison with Fig. 4a suggests that the SCZ curve approximates an SNR of 0.2 in our simulations, whereas disorders like ASD have very low SNR (i.e., SNR∼0.05) indicating that extremely large samples would be required to achieve consistent findings (see 8 in Online Methods).

## Discussion

A common assumption in biological psychiatry is that robust neurobiological phenotypes for psychiatric disorders, as defined using current diagnostic systems, can be identified once enough well-designed studies have been conducted. However, several decades of structural MRI research in psychiatry have thus far failed to achieve such convergence on robust patterns of grey matter thickness or volume difference. Our analysis, which focuses on two of the most widely-studied biological phenotypes––MRI-based measures of differences in grey matter thickness and volume––indicates that current practice in structural MRI research, which heavily relies on multiple independent studies of small samples, is unlikely to ever converge on robust findings, given that average CSCs never exceed *r* = 0.16.

Low cross-site consistency could be attributable to: (1) between-study variations in processing and analysis methods; (2) between-study variations in the clinical and/or demographic characteristics of the samples studied; (3) a reliance on small sample sizes; (4) fundamental problems with the diagnostic constructs themselves; and/or (5) fundamental limitations of structural MRI morphometry for detecting robust, clinically actionable biological phenotypes of psychiatric disease. Our analyses indicate that the first two factors do not make a major contribution to cross-site consistency, since CSC estimates were low across a wide range of processing and analysis pipeline choices related to statistical thresholding, data smoothing, and regional parcellation and were also generally unrelated to cross-site variations in the numerous clinical and demographic characteristics for which data were available (barring some specific exceptions). However, our analysis was necessarily limited to summary statistics and could not examine more fine-grained details, such as differences in the specific class of medication administered to cases or variations in the severity of specific symptom dimensions.

Our bootstrap analyses indicated that the third factor, sample size, influences CSC in AD, SCZ, and SCA. SCA showed a similar trend to SCZ, which may reflect broader phenotypic similarities between the two conditions^58^. Our findings suggest that sample sizes beyond *N*∼200 will start to converge on a consistent morphometric phenotype for SCZ, but that samples above 400 are likely required to approach the level of consistency seen in AD at *N*∼60. This effect of sample size suggests that schizophrenia-spectrum disorders are indeed associated with an identifiable morphometric phenotype, albeit one that is not observed as strongly as in AD. Whether this difference represents a stronger neuropathological substrate for AD, or the fact that the AD patients from different sites were scanned at a more similar illness stage within our dataset, remains unclear, although between-site differences in sample age were generally not significantly related to CSCs in psychiatric disorders (Fig. 3). Either way, our results showed that increasing sample sizes in studies of SCZ will facilitate suppression of inter-individual variability and between-study variations in methodological, demographic, and clinical characteristics to support robust delineation of the underlying disease effect. Pooling data from multiple sites is the most cost-efficient strategy for achieving this goal, with consortia such as the Enhancing Neuroimaging Genetics through Meta-Analysis (ENIGMA) consortium being one successful example of such an approach^59,60^. The data analyzed in such consortia are generally limited to region-level measures obtained with coarse parcellations. Repositories for storing and pooling voxel-level data would facilitate robust phenotype identification.

For other disorders, we found less evidence that moderate increases in sample sizes will improve cross-site consistency, although we could not examine samples exceeding 150-250 people per group. Comparison with our simple simulations suggests that the disease phenotype SNR for these disorders is much lower than in SCZ, which is consistent with empirical analyses suggesting that SCZ patients show more extensive grey matter reductions than people diagnosed with other disorders^34,61^. Indeed, our simulations indicate that sample sizes in the order of 1000 may be required to obtain CSCs of *r* > 0.5 for MDD, ASD, and BD, which were obtained with N=30 for AD and N=200 for schizophrenia. This result is in line with recent work showing that the CSC for depression-related cortical thickness difference maps obtained in the UK Biobank (N=2220 cases; 2590 controls) and through the ENIGMA consortium (2148 cases, 7957 controls), parcellated with the DK atlas, is *r* = 0.64^62^. This result, combined with our findings, is consistent with a disease phenotype SNR=0.05 in our simulations (Extended Data Fig. 10b). Thus, while meta-analyses may be able to reveal statistically significant differences by pooling datasets to obtain sufficiently large samples^8,63,64^, the consistency of any pair of individual studies will be low if sample sizes remain at current levels. This low consistency means that any attempt to compare or relate the results of one small-sample study to another, which is commonplace in the field, is essentially meaningless. The comparatively low SNR of the disease phenotype in MDD, ASD, and BD may reflect a higher degree of phenotypic heterogeneity in these conditions or may arise from a potential ascertainment bias, in which people with SCZ are more likely to come from tertiary referral centers and therefore experience a more severe form of illness^65,66^.

The low consistency we observed here aligns with an increasing realization that current diagnostic constructs likely refer to multiple disorders that arise from distinct causes^2,67,68^ and that grey matter alterations occurring at the level of individual patients with the same diagnosis occur in highly heterogeneous locations, suggesting that group means do not accurately reflect the changes occurring within individuals^33–36^. The definition of more homogeneous diagnostic constructs or subgroups may thus be needed to augment the SNR of the underlying disease phenotype^69^. Approaches to characterize psychiatric phenotypes using clinical or other measures, such as identification of genetic sub-types (e.g, 22q11 deletion and other psychiatric syndromes resulting from rare genetic mutations^70^), the Research Domain Criteria (RDoC)^71^, and the Hierarchical Taxonomy of Psychopathology (HiTOP)^72^ frameworks, may be helpful in this regard^2^. Other approaches include the use of unsupervised clustering^73–77^, multivariate^78,79^, and network-based approaches^34,80–84^ to better identify commonalities between individual cases than classical mean-based SBM and VBM comparisons.

The low consistency observed for structural-MRI-derived grey matter differences may not be specific to psychiatric disorders. Even though our findings suggest that structural MRI can identify a robust morphometric phenotype for AD, inconsistencies have been noted in relation to conditions such as epilepsy and migraine^85,86^, potentially reflecting their aetiological heterogeneity or the limitations of MRI-based morphometry. Different imaging modalities may be more effective at revealing consistent disease phenotypes^87^. For instance, positron emission tomography (PET) imaging using FDOPA has consistently shown frontostriatal dopamine dysregulation across different stages of psychotic illness^88–90^ and may differentiate cases who respond to antipsychotics from those who do not^91^. In depression, functional MRI-derived measures of brain activity show stronger effect sizes than those obtained with cortical thickness^62^. The approach developed here could be used to benchmark which imaging modalities and measures are able to yield the most robust biological phenotypes for different disorders.

Although it is widely appreciated that large samples are required for robust inference^21,92^, the extent of the problem posed by reliance on small samples in psychiatric neuroimaging, a field littered with inconsistent findings, has not been previously quantified. Our analysis, which explicitly tries to quantify the problem, indicates that current practices, in which multiple investigators report on the results of case-control comparisons conducted in samples of less than 100 people per group, are unlikely to ever converge on a robust morphometric phenotype for psychiatric disorders when using structural MRI. Instead, future efforts should prioritize (1) transdiagnostic and consortia-driven initiatives (e.g., UK Biobank^93^, Chinese Color Nest Project (CCNP)^94^, German National Cohort^95^, IMAGEN study^96^, ENIGMA^59^) and the open release of data to establish large repositories for robust biomarker identification; (2) the use of harmonized, transdiagnostic clinical assessment batteries to facilitate the identification of more homogeneous disease subgroups^2,97^; and (3) the investigation of approaches other than mass univariate analysis of grey matter volume or cortical thickness for disease phenotyping. Furthermore, analyses that better account for inter-individual heterogeneity^2,34^ may facilitate robust localization of pathophysiological markers in homogeneous patient sub-groups, facilitating target identification for brain stimulation and other treatments.

## Data Availability

All data produced in the present study are available upon reasonable request to the authors

## Acknowledgements

This work was supported by the MASSIVE HCP facility (http://www.massive.org.au). This research was supported by the following grants: Australian National Health and Medical Research Council (ID: 2034000), JP; Monash FMNHS Early Career Research Excellence Program, JP; Australian Government Research Training Program Scholarship, MG; European Research Council (ERC) under the European Union’s Horizon 2020 research and innovation programme (866533-CORTIGRAD), AH; The University of Melbourne McKenzie Fellowship, SC; The Brain & Behaviour Research Foundation Young Investigator Grant, SC; NHMRC Investigator Grant 2033976, JT; Sylvia and Charles Viertel Charitable Foundation (Viertel Charitable Foundation), AF; National Health and Medical Research Council (IDs: 1146292 and 1197431), AF; Australian Research Council (IDs: DP200103509 and FL220100184), AF. The funders had no role in study design, data collection and analysis, decision to publish or preparation of the manuscript. We thank all the sites and investigators who have worked to share their data through data sharing repositories including ABIDE, AIBL, NDA, SchizConnect, 1000 Functional Connectomes Project, MACC, MBBP, OASIS, SRPBS, MIRIAD, YoDA, and OpenNeuro. Finally, we would like to thank … and the anonymous reviewer who provided valuable feedback.

## Author Contributions Statement

TC, JP, and AF designed and revised the study and interpreted the results. TC collated, preprocessed, and analysed data and wrote the manuscript. MG reviewed the code for data analysis. JW contributed to the data analysis. MG, AS, AH, SC, JHZ, FJ, and TCo contributed to data preprocessing. JHZ, CC, FJ, BH, CD, JT, BH, JK, MB, and AF contributed to the data collection and acquisition. JP and AF critically revised the manuscript. All authors reviewed the manuscript and approved the decision to submit for publication.

## Methods

### 1. Data curation

To examine the consistency of grey matter differences between people with psychiatric disorders and controls, we collated data from numerous open repositories and locally acquired data to which we had access. Open repositories were identified by a general search as well as searches within large repositories. We retained datasets if they met the quality control described in **Error! Reference source not found.** and **Error! Reference source not found.**. A complete list of included datasets is provided in Supplementary Table 1. Each dataset was acquired in accordance with local ethics committee regulations, as detailed in the original publications (see Supplementary Table 1). The present analyses were approved by the Monash University Ethics Committee. Our curation procedure resulted in a total pool of 133 studies with 165 independent scan sites, where we considered data from one scanner as an individual site (a single study may therefore contribute multiple ‘sites’ in our parlance if it included data from multiple scanners). We initially included attention deficit hyperactivity disorder, obsessive-compulsive disorder, and anxiety in the search of literature and data, but we could not obtain sufficient data to allow robust analysis.

### 2. Data quality control

The experimental design and quality controls used at each site have been described in the corresponding articles (see Supplementary Table 1 for references). We only retained data from participants from 18 to 60 years of age in each site (except for sites for Alzheimer’s disease) and where T1-weighted MRI and sufficient clinical or demographic data (e.g., indication of diagnosis, age, sex, or scanner site) were available. We further excluded sites with fewer than 20 participants per diagnostic group. We chose the cut-off of 20 participants as it is a common sample size in the field and smaller samples are likely to yield very unreliable findings, further reducing CSCs (see Fig. 3 in ^56^). After preprocessing, we performed additional quality control checks and excluded participants with images that did not successfully complete our preprocessing pipeline or with low-quality image outputs according to the specific criteria outlined below. Sites with fewer than 20 participants per group remaining after these procedures were also excluded. Due to variations in the data quality checks for SBM and VBM, the two analyses examined different numbers of participants, as detailed below.

### 3. Measuring grey matter differences

We quantified group differences in cortical thickness and grey matter volume using SBM and VBM, respectively, because they are the two most widely used methods for characterizing morphometric alterations in psychiatric disorders. Data from each site were processed and case-control group differences in cortical thickness or grey matter volume were quantified using the same analysis approach, as discussed below. In reality, different studies run by different investigators rely on different processing and analysis pipelines, which attenuates cross-site consistency. Our use of a unified protocol, coupled with our correction for site effects (see below), means that our analysis provides an optimistic upper bound on the level of cross-site consistency that can be attained.

#### 3.1 Surface-based morphometry (SBM)

##### Quality Control Procedures

Image quality control (QC) was performed through a combination of visual inspection of surface reconstruction (from FreeSurfer^38,39^) accuracy, auto-detection of image quality outliers with MRIQC^98^, and analysis of Euler numbers^38,99^ (a widely used measure of image quality). In particular, we used the MRIQC software package^98^ to extract 42 different measures quantifying various aspects of image quality. We applied principal component analysis (PCA) on these measures to extract the first *k* dimensions accounting for 80% of the variance and identified participants with a normalized score exceeding z = 3.5 (i.e., 99.98 percentile) on any of the *k* principal components as outliers. The PCA was run separately for each site, and so the specific value of *k* was site-specific. Outlying participants were manually inspected for obvious image artifacts or surface reconstruction inaccuracies, which resulted in participant exclusion. We also used Euler numbers calculated from FreeSurfer and excluded participants that had averaged Euler numbers (across two hemispheres) smaller than 3.29 standard deviations below the sample mean^38,99^. After participant-level exclusions, we excluded sites with less than 20 participants per group, yielding a final data pool of 3091 cases (AD and psychiatric disorders) and 3002 controls from 66 sites for the SBM analysis.

##### SBM-based estimation of cortical thickness

We estimated cortical thickness using SBM of the T1-weighted MRI scans, as implemented in FreeSurfer version 7.1.0^38,39^. The FreeSurfer SBM pipeline includes: volumetric registration to MNI305 atlas^100^; bias field estimation and removal; removal of non-brain tissue using a deformable template model^101^; tissue and broad brain region segmentation using a combination of intensity and atlas-based approaches^102^; generation and refinement of a triangular surface mesh model of the white matter surface by following the intensity gradients between the white and grey matter; generation of a pial surface model by expanding the white matter surface to follow intensity gradients between the grey matter and cerebrospinal fluid (CSF); and then estimation of cortical thickness at each surface vertex as the average distance between the closest points on the white and the pial surfaces^103^. The pial surface was inflated and aligned via spherical registration to the *fsaverage* atlas, which comprises 163,842 vertices per hemisphere^104^. The cortical thickness measures were then smoothed using a surface-based Gaussian kernel with a full-width at half maximum (FWHM) of 10 (results reported in the main text), 15, and 20 mm.

##### Adjusting for site effects

We adjusted each participant’s cortical thickness map (SBM) for site-specific effects using the data harmonization method, ComBat^54^. ComBat models site-specific scaling factors and uses empirical Bayes to improve the estimation of the site parameters and to preserve biological variability of interest while removing systematic differences between sites. We ran the model using the publicly available neuroCombat package in Python (https://github.com/Jfortin1/neuroCombat), specifying site as the batch variable and including age, sex, and diagnosis as covariates to preserve clinically relevant variance. ComBat was applied to minimize between-site differences, but it may result in an optimistic assessment of cross-site consistency or potentially remove site-specific group differences. The results presented in the main text (i.e., Fig. 2) were generated using ComBat-corrected data. We also re-ran the analyses without ComBat.

##### Statistical analysis

After adjusting each thickness map for scan site effects with ComBat^54^, we ran general linear models using the *mri_glmfit* command on each hemisphere in FreeSurfer, taking into account sex and age as nuisance covariates. We obtained similar results when including Euler number (a measure of image quality^38^) or total intracranial volume (ICV) as a covariate in addition to age and sex (Supplementary Fig. 9). The model results in *F*-statistic and *p*-value maps, which were then used to generate *z*-statistic maps quantifying the magnitude of case-control differences in cortical thickness at each vertex. Thresholded maps were obtained by thresholding the *p*-value maps at two different thresholds: *p* < 0.05, uncorrected, and *p* < 0.05, corrected. The corrected threshold was estimated using a commonly-used cluster-wise correction procedure in which clusters of contiguous vertices showing statistically significant effects are identified and their significance evaluated using permutation testing, as implemented in the *mri_glmfit-sim* command^105^.

##### Region-level analyses

To evaluate whether region-based analyses result in a higher level of cross-site consistency, we parcellated each participant’s unsmoothed cortical thickness maps using the Desikan-Killiany^52^ and Schaefer-100, Schaefer-500, and Schaefer-1000 atlases^53^, resulting in 34, 50, 250, and 500 regions per hemisphere. Note: the 500-region right hemisphere atlas only comprises 498 regions as a result of the generating algorithm. The mean cortical thickness of each parcel was extracted and used as an input for the same analysis described above.

#### 3.2 Voxel-based morphometry (VBM)

##### Quality control procedures

QC was performed through a combination of visual inspection of grey matter segmentation and spatial normalization accuracy on native and Montreal Neurological Institute (MNI) spaces and analysis of image quality ratings (IQR) extracted from Computational Anatomy Toolbox (CAT12)^106^. In particular, we quantified grey matter volume using VBM of the T1-weighted MRI scans, as implemented in CAT12 (version 8.1, http://dbm.neuro.uni-jena.de/cat/)^106^, which is included in the Statistical Parametric Mapping software (SPM12, http://www.fil.ion.ucl.ac.uk/spm/software/spm12)^107^ in MATLAB r2021a. As an initial step, we extracted weighted IQR from CAT12 preprocessing reports and excluded participants with IQR>2.8. The IQR combines estimates of noise-to-contrast ratio, inhomogeneity contrast ratio, and root mean square resolution^106^. IQR ranges from 0.5 (excellent) to 10.5 (unacceptable) to indicate image quality. The threshold for IQR was chosen based on previous studies^35^. We excluded participants based on their IQR score and sites that had less than 20 participants per group after IQR-based exclusion, yielding a total of 2427 cases and 2353 controls from 42 sites for the VBM analysis. We used the IQR-based approach for VBM to emulate procedures typically used by published in the field^108^.

##### Data processing and volume estimation

The CAT12 pipeline includes the following steps: spatial adaptive non-local means (SANLM) denoising filter^109^, resampling, bias correction, affine registration, initial unified segmentation^110^, refined segmentation and skull-stripping, local intensity correction, adaptive maximum a posteriori (AMAP) segmentation^111^, partial volume estimation^112^, and spatial normalization to MNI space using the MNI152NLin2009cAsym template by Geodesic Shooting^113^. We applied volumetric Gaussian smoothing with kernels sizes (FWHM) of 6, 8, and 12 mm to the preprocessed maps to improve signal-to-noise ratio. To constrain our analyses to grey matter voxels, a common mask was generated from all the normalized, modulated, warped grey matter maps retaining voxels with a tissue probability ≥ 0.2^114^. We created a common mask from all psychiatric sites to account for additional atrophy expected to occur in the older AD samples.

##### Statistical analysis

Site effects were adjusted using the same ComBat^54^ model applied to the cortical thickness maps. We then quantified group differences in grey matter volume using voxel-wise general linear models, including total intracranial volume, sex, and age as covariates, and estimated *t*-statistics within the common mask to quantify the magnitude of voxel-level control-case differences in grey matter volume for each site. Corresponding *p*-values of the *t*-statistics were estimated via random field theory^115,116^. The uncorrected thresholded maps (controls>cases and controls<cases) were obtained by applying a voxel-wise threshold of *p* < 0.05. The corrected thresholded maps were obtained using family-wise error rate (FWER) correction at *p* < 0.05 at the cluster level, with an initial height threshold of 0.05 and extent threshold of 0 voxels.

##### Region-level analyses

Mirroring the SBM analysis, we also parcellated the unsmoothed grey matter maps of each participant in MNI space (mwp1 – no smoothing) using the Schaefer-100, Schaefer-500, and Schaefer-1000 region parcellations (7-network version)^53^, combined with the Melbourne subcortical parcellation (32 regions)^117^ and Buckner cerebellar parcellation (7 regions)^118^. The grey matter volume of each region was then quantified as the mean intensity of all voxels in the regions. All analyses were repeated using these regional volume estimates.

### 4 Quantifying cross-site consistency

The SBM and VBM analyses result in a site-specific map of case-control differences in cortical thickness and grey matter volume, respectively. To examine the consistency of the resulting spatial patterns across sites, we first calculated the product-moment correlation (Pearson’s), *r*, of the unthresholded statistical maps between each pair of sites, separately for each disorder. This procedure resulted in a site × site matrix of cross-site consistency estimates for each disorder (see Figs. 1, 2, and Extended Data Fig. 2). Note that since we are interested in correlations of the spatial profile the results do not change if using the *z*-statistics or the *t*-statistics, estimated as default statistics in FreeSurfer or SPM12, respectively. As Freesurfer pipelines and analyses are applied to each hemisphere independently, we examined CSCs for SBM separately in the left and right hemispheres.

For thresholded maps (both uncorrected and corrected), we quantified cross-site consistency using binary correlations^49,50^ and replication rates^21,51^ after converting the maps into one-dimensional vectors. The binary correlation of two vectors was quantified by the mean square contingency coefficient 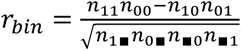, where *n*_*xy*_, with *x*, *y* = {0,1}, counts the number of pairs of values observed together from the same element of each vector. For example, *n*_10_ counts the number of times that the same element in the first vector is equal to 1 (i.e., x = 1) and in the second vector is equal to 0 (i.e., y = 0). The symbol ∎ represents that the element is either 1 and 0 (i.e., *n*_1∎_ = *n*_11_ + *n*_10_). The replication rate, *RR*, introduced in prior work^21,51^, was quantified as the number of times a value of 1 is observed in both vectors normalized by the number of 1s observed in one vector, 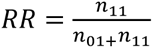.

### 5 Statistical inference on cross-site consistency

#### 5.1 Null models for SBM

To evaluate whether the magnitudes of the spatial correlations between site-specific cortical thickness difference maps were consistent with chance expectations, we used eigenstrapping^42^ to generate an ensemble of randomized spatial maps with the same autocorrelation as the original data. Eigenstrapping involves a spectral decomposition of the cortical thickness difference map obtained for each site using geometric eigenmodes as a basis^119^. The eigenmodes were randomly rotated and used to generate a new map that preserves the original spatial autocorrelation but randomizes its other statistical features. The procedure results in better false-positive control than commonly used methods, such as the spin test, while avoiding problems posed by medial wall vertices (see ^42^ for a detailed discussion).

Each site-specific map was subjected to this procedure 1000 times to generate surrogate *z*-statistic maps, thresholded maps, and corrected thresholded maps for each smoothing kernel (0, 10, 15, and 20 mm) on the left hemisphere. We also ran the analyses for the right hemisphere with a 10 mm smoothing kernel. The surrogate thresholded maps were obtained by retaining the *V* vertices with the highest values, where *V* is the number of vertices that survived the (uncorrected or corrected) thresholding procedure applied to the empirical data. For each iteration and each type of map, we estimated the null cross-site consistency matrix to generate an empirical null distribution representing the magnitudes of consistency estimates expected by correlating randomized maps with the same spatial autocorrelation as the original data. The median of these matrices was then extracted to form a null distribution for inference on the observed median cross-site consistency value. For region-level analyses, we parcellated the vertex-wise null *z*-statistic maps generated from the *z*-statistic maps of unsmoothed CT maps on DK and Schaefer-100, Schaefer-500, and Schaefer-1000 atlases and repeated the above procedures to generate the null ensembles.

We also considered a permutation-based null model. This model involved permuting the labels of the patients and controls 100 times and then repeated the statistical analysis to obtain the null group difference maps. At each permutation, we correlated the null difference maps between pairs of sites to obtain a null CSC matrix and the median correlation was extracted from this matrix at each iteration to construct a null distribution for the observed median cross-site consistency value. (Supplementary Fig. 1 and 2).

Note that the SBM null distribution is shifted towards higher CSCs in AD relative to other disorders likely because the widespread anatomical distribution of AD case-control differences increases the autocorrelation of the spatial map, which in turn inflates correlations between null maps with matched autocorrelation.

#### 5.2 Null models for VBM

Eigenstrapping is an excellent inferential procedure for spatial correlations between surface maps^42^ but does not generalize to whole-brain volumetric images. For VBM, we generated null ensembles using BrainSMASH^43^(Brain Surrogate Maps with Autocorrelated Spatial Heterogeneity), which randomly permutes the map values, smooths, and rescales the permuted map to best match the spatial autocorrelation of the original map. For each site-specific grey matter volume difference map processed with a 6 mm smoothing kernel, we generated 1000 surrogate *t*-statistic maps using this procedure. Due to the computational expense of BrainSMASH, we only generated null ensembles for this smoothing kernel and used them as an approximate null benchmark for results generated with 8 mm and 12 mm kernels and results without ComBat. The same procedure used in SBM was applied to the BrainSMASH nulls to generate surrogate data for thresholded and region-level analyses. We also performed permutation-based null tests for VBM as in SBM. Note that the permutation nulls preserve the sample size, variance, and spatial autocorrelation of the input data at each site, while the BrainSMASH and eigenstrapping nulls were generated to more directly match the spatial autocorrelation with that of the observed group difference map, which was used for CSC calculation.

### 6 Quantifying the effects of sample and MRI data characteristics

We evaluated whether 19 different clinical, demographic, and data characteristics (Table 2) were related to CSC values across different pairs of sites, thus allowing us to quantify whether site pairs with more similar grey matter difference maps were more closely matched in terms of these characteristics. For each site, we extracted a single scalar value summarizing the site’s average or total (as appropriate) for that characteristic and then computed a ratio of scalar values for each pair of sites, resulting in a site-by-site matrix of similarity estimates for each characteristic. In other words, for any given characteristic, *C*, estimated for sites *i* and *j*, we defined a ratio matrix as

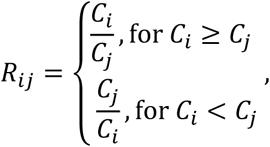

with *R*_*ij*_ = 1 indicating equivalence between sites and *R*_*ij*_ > 1 indicating an imbalance in one site relative to the other. The ratios we examined were: (1) mean participant age; (2) variance of participant ages; (3) number of male participants; (4) number of female participants; (5) male-to-female ratio of participants; (6) number of cases; (7) number of controls; (8) total number of participants; (9) case-to-control ratio; (10) medication exposure, which was quantified using a binary indicator based on whether people with psychiatric disorders were taking antipsychotics, mood stabilisers, antidepressants, or anxiolytics at the time of scanning; (11) mean Euler number of cortical surface reconstructions; (12) variance of the Euler numbers; (13) mean of age of illness onset; (14) variance in the age of illness onset; (15) mean illness duration; (16) variance of illness duration; (17) MRI scanner manufacturer; (18) MRI scanner model; and (19) MRI voxel size. For discrete values such as medication exposure, MRI scanner manufacturer, and MRI scanner model, we compared sites using a binary indicator (i.e., *R*_*ij*_ = 1 for similar; *R*_*ij*_ = 0 otherwise). In the VBM analyses, we considered similar site properties except that we used the IQR mean and IQR variance to represent image quality instead of Euler number mean and Euler number variance.

We then used Spearman rank correlations to quantify the association between each independent element of the cross-site characteristic similarity matrix and the corresponding element of the cross-site consistency matrix for each disorder. Statistical inference on these correlations was performed using Mantel tests^55^, which correlates the upper triangles of the two matrices and obtains *p*-values via a permutation procedure that accounts for the symmetry of the matrices.

### 7 Examining the effects of sample size

We used bootstrapping to examine how sample size influences cross-site consistency. First, we combined data from all cases and all controls into a single consolidated patient population and a single control population for each disorder. Second, from the case population, we randomly sampled two case groups of the same sample size. Different sample sizes were chosen to be equally spaced on a logarithmic scale and with the maximum sample size corresponding to approximately half the total number of participants in case group in the pooled dataset. For example, the total available pool of cases was 1054 for schizophrenia, so the sample sizes were *N* = {10, 16, 25, 40, 63, 100, 158, 251, 398, 527}, with 527=1054/2 being the largest sample size. Third, we randomly selected two groups of controls with sexes and ages that best matched the selected cases. Each pair of case and control sub-groups formed a pseudo-site. Fourth, we computed two *z*-maps of cortical thickness difference––one for each pseudo-site––and correlated them. We repeated these four steps 100 times, allowing us to obtain a distribution of cross-site consistency estimates for a given sample size. We repeated the procedure for cortical thickness maps parcellated using the DK, Schaefer-100, Schaefer-500, and Schaefer-1000 atlases and examined the consistency of correlations between unthresholded maps, binary correlations between thresholded maps, and replication rates of thresholded maps.

This analysis may provide an optimistic estimate of the cross-site consistency observable for any given sample size, since (a) all images were processed and analyzed with a uniform protocol, which does not reflect the reality in the literature; (b) the data were preprocessed with ComBat to remove residual site-specific effects (although our main analysis indicates that this step did not appreciably impact empirical cross-site consistency estimates); and (c) we only considered balanced sample sizes of cases and controls. However, our total data pool for each case group was derived by combining multiple heterogeneous studies recruiting cases at different illness stages and with demographic characteristics, which may reduce consistency. Our bootstrapping analysis thus offers a coarse approximation of the sample size required to achieve a particular level of cross-site consistency.

### 8 Simulating the effects of sample size

As the number of participants available for each disorder varied considerably and limited our investigation of large samples in the bootstrapping analysis, we used a simple simulation to examine how sample size impacts consistency under different assumptions about the degree to which a common disease phenotype is expressed on the brain by individuals with the same disorder, above and beyond intrinsic variability. We first defined a synthetic cortical thickness map for each individual by generating vectors of length 100,000 (Fig. 4b) or 1000 (Extended Data Fig. 10), representing individual vertices or regions of interest, by randomly sampling from Gaussian distribution with mean = 0 and standard deviation = 1. For each individual in the patient group, we added a common random vector of the same length, representing a shared disease phenotype, with a contribution that is tuned by a scalar parameter *α*. Formally, we have

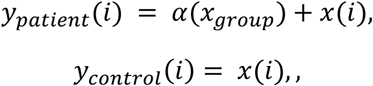

where *x*_*group*_ = *N*(0,1) is a random vector common to all individuals in the patient group, thus representing a common disease phenotype, and *x*(*i*) = *N*(0,1) is a random vector unique to each individual, thus representing inter-individual variability and/or measurement noise. The parameter *α* thus tunes the SNR of the disease phenotype relative to random individual variation. We caution that comparisons between bootstrap analysis and simulation results are only for illustrative purposes and our model is too simplistic to directly infer the SNR of each disease phenotype.

For each simulation, we generated maps for individuals from control and case groups with sample sizes for each group ranging from 10 to 10000. At each sample size, we used the same procedure as in our boostrapping analysis, in which we generated two pseudo-sites and quantified their cross-site consistency, repeating the process 5 times at each sample size, thus corresponding to 5 different pairs of sites (Fig. 4b and Extended Data Fig. 10a and b). We also examined the impact of varying the number of study sites by changing the repetition at each sample size to 50 times (Extended Data Fig. 10c). Adding more sites with the same sample (Extended Data Fig. 10a and c) or changing the vector (map) lengths (Fig. 4b and Extended Data Fig. 10) does not change the mean but does reduce the variance across repeated experiments.

### Data Availability

Data access methods are specified in Supplementary Table 1, which we re-produce here

Autism Brain Imaging Data Exchange I (ABIDE I), available in the ABIDE I repository, http://fcon_1000.projects.nitrc.org/indi/abide/abide_I.html

Autism Brain Imaging Data Exchange (ABIDE II), available in the ABIDE II repository, http://fcon_1000.projects.nitrc.org/indi/abide/abide_II.html

Australian Imaging, Biomarker & Lifestyle (AIBL), available in the AIBL repository, subject to the approval of the AIBL Management Committee https://aibl.org.au/, https://ida.loni.usc.edu/

Advancing innovative brain imaging to detect altered glutamate modulation and network dynamics in schizophrenia (AIS*), available on NDA, subject to the approval of Data Access Committee, https://nda.nih.gov/edit_collection.html?id=2488

Bipolar & Schizophrenia Consortium for Parsing Intermediate Phenotypes (B-SNIP 1), available on NDA, subject to the approval of Data Access Committee, https://nda.nih.gov/edit_collection.html?id=2274

Bipolar & Schizophrenia Consortium for Parsing Intermediate Phenotypes (B-SNIP 2), available on NDA, subject to the approval of Data Access Committee, https://nda.nih.gov/edit_collection.html?id=2165

The Autistic Brain Over 45: The Anatomic, Functional, and Cognitive Phenotype(ASD45*), available on NDA, subject to the approval of Data Access Committee, https://nda.nih.gov/edit_collection.html?id=2291

Glutamatergic and Neuronal Dysfunction in Gray and White Matter (BrainGluSchi), available in the BrainGluSchi repository, http://schizconnect.org/

The Center for Biomedical Research Excellence (COBRE), available in the COBRE repository, http://fcon_1000.projects.nitrc.org/indi/retro/cobre.html

The Determinants of Social Disconnectedness: A Spectrum from the General Community to Severe Mental Illness(DSD*), available on NDA, subject to the approval of Data Access Committee, https://nda.nih.gov/edit_collection.html?id=2481

Human Connectome Project for Early Psychosis (HCP-EP), available on NDA, subject to the approval of Data Access Committee, https://www.humanconnectome.org/study/human-connectome-project-for-early-psychosis, https://nda.nih.gov/edit_collection.html?id=2914

Singapore Memory Ageing and Cognition Centre (MACC) Harmonization Project (Harmonization), available from the principal investigator of the study, subject to local ethics committee requirements

Inhibitory dysfunction in autism (IDAUT*), available on NDA, subject to the approval of Data Access Committee, https://nda.nih.gov/edit_collection.html?id=2266

Monash Brain and Behaviour Project (MBBP), available from the principal investigator of the study, subject to local ethics committee requirements

MIND Clinical Imaging Consortium (MCIC), available in the MCIC repository, http://schizconnect.org/

Modulation of ventrolateral prefrontal cortical activity during reward processing by transcranial direct current stimulation (MRITDCS*), available on NDA, subject to the approval of Data Access Committee, https://nda.nih.gov/edit_collection.html?id=2432

A Multidimensional Investigation of Cognitive Control Deficits in Psychopathology (MICCD*), available on NDA, subject to the approval of Data Access Committee, https://nda.nih.gov/edit_collection.html?id=2102

Cortical myelin measured by the T1w/T2w ratio in individuals with depressive disorders and healthy controls (Myelin*), available in the repository, https://openneuro.org/datasets/ds003653/versions/1.0.0

Open Access Series of Imaging Studies (OASIS-3), available on OASIS repository, subject to the approval of Knight ADRC, https://www.oasis-brains.org

Open Access Series of Imaging Studies (OASIS-4), available on OASIS repository, subject to the approval of Knight ADRC, https://www.oasis-brains.org

Psychosis and Affective Research Domains and Intermediate Phenotypes (PARDIP), available on NDA, subject to the approval of Data Access Committee, https://nda.nih.gov/edit_collection.html?id=2126

Resting state with closed eyes for patients with depression and healthy participants (RD*), available in the repository, https://openneuro.org/datasets/ds002748/versions/1.0.5

Japanese Strategic Research Program for the Promotion of Brain Science (SRPBS), available in the SRPBS repository, subject to the approval of the DecNef Consortium, https://bicr.atr.jp/decnefpro/data/

Specificity of Hippocampal Subregion Cerebral Blood Volume Abnormalities in Psychiatric Disorders (SHSCBV*), available on NDA, subject to the approval of Data Access Committee, https://nda.nih.gov/edit_collection.html?id=3199

A Study of the neural and psychological correlates of emotion and motivation processing deficits in individuals with schizophrenia vs. controls (EMOSCZ*), available on NDA, subject to the approval of Data Access Committee, https://nda.nih.gov/edit_collection.html?id=2703

The Transdiagnostic Connectome Project (TCP), available in the repository, https://openneuro.org/datasets/ds005237

UCLA Consortium for Neuropsychiatric Phenomics (UCLA-CNP), available in the UCLA repository, https://openneuro.org/datasets/ds000030/versions/00016

Youth Depression Alleviation-Combined Treatment (YoDA), available from the principal investigator of the study, subject to local ethics committee requirements

Minimal Interval Resonance Imaging in Alzheimer’s Disease (MIRIAD), available in the MIRIAD repository, https://www.ucl.ac.uk/drc/research-clinical-trials/minimal-interval-resonance-imaging-alzheimers-disease-miriad

Brain correlates of speech perception in schizophrenia patients with and without auditory hallucinations (Speech*), available in the repository, https://openneuro.org/datasets/ds004302/versions/1.0.0

### Code Availability

Data were preprocessed with FreeSurfer (version 7.1.0, https://surfer.nmr.mgh.harvard.edu/) and Computational Anatomy Toolbox (CAT12, version 8.1, http://dbm.neuro.uni-jena.de/cat/), which is included in the Statistical Parametric Mapping software (SPM12, http://www.fil.ion.ucl.ac.uk/spm/software/spm12). Data were harmonised to reduce site effects by neuroCombat package in Python (https://github.com/Jfortin1/neuroCombat). Analysis and figures were performed using MATLAB R2023b. All code used to produce the results and figures for this study can be found at https://github.com/NSBLab/reproducibility_grey_matter_differences_in_disorders.

